# Temporal Patterns and Risk Factors of Diarrheal Comorbidity among Children aged < 5 years in Rural Western Kenya: Evidence from Three Consecutive Enteric Studies─2008-2024

**DOI:** 10.1101/2025.11.11.25340031

**Authors:** Billy Ogwel, Bryan O. Nyawanda, Brian O. Onyando, Alex O. Awuor, Caleb Okonji, Raphael O. Anyango, Caren Oreso, Catherine Sonye, John B. Ochieng, Stephen Munga, Dilruba Nasrin, Karen L. Kotloff, Patricia B. Pavlinac, Richard Omore, Elizabeth T Rogawski McQuade

## Abstract

Sub-Saharan Africa bears the highest burden of diarrhea, often complicated by comorbidities that delay diagnosis, hinder treatment, and worsen outcomes. As the epidemiology of diarrheal disease evolves, understanding comorbidity patterns is critical for effective public health responses. We examined the temporal patterns and risk factors of diarrheal comorbidity in Kenyan children aged < 5.

We conducted secondary pooled analysis with a retrospective cohort design leveraging data from the Global Enteric Multicenter Study (GEMS, 2008-2012), the Vaccine Impact on Diarrhea in Africa (VIDA, 2015-2018), the Enteric for Global Health (EFGH) Shigella surveillance study (2022-2024). The outcome was comorbidity count, defined by Integrated Management of Childhood Illnesses case definitions and clinician diagnoses of ten conditions: malaria, bacterial infection, pneumonia, severe acute malnutrition (SAM), meningitis, acute febrile illness (AFI), respiratory Illness (non-pneumonia), anemia, stunting and wasting. Temporal trends were assessed using descriptive statistics and the Cochran-Armitage trend test. Risk factors were identified using generalized estimating equations with a Poisson distribution, adjusting for clustering. We analyzed data from 4,148 children with moderate-to-severe diarrhea; 90.3% had ≥ one comorbidity, with a declining trend across studies: GEMS (92.9%), VIDA (89.3%), and EFGH (86.6%). Pneumonia (49.5%), malaria (48.3%), and stunting (24.7%) were most common comorbidities. The proportion of children with only one comorbidity increased (28.9% [2008] to 49.7% [2024]), while multiple comorbidities declined. Traditional comorbidities (malaria, pneumonia, wasting, SAM) significantly decreased, while AFI, anemia, and non-pneumonia respiratory illness increased. Multivariable analysis identified older age, lower caregiver education, dehydration, vomiting, and high respiratory rate as drivers of higher comorbidity counts, while female sex was associated with fewer comorbidities.

Despite the high prevalence, we observed a 25–29% decline in comorbidity burden and a fundamental shift in disease profiles. Our findings support the need for a shift from single-disease control to integrated disease management.

## Introduction

While diarrheal deaths have decreased, diarrhea remains a major public health problem, disproportionately affecting sub-Saharan Africa (1). This challenge is amplified by comorbidities, co-occurring conditions, which complicate the clinical course of an index disease (2). The presence of comorbidities increases disease complexity and severity, hinders diagnosis, delays appropriate treatment, and elevates the risk of adverse outcomes and associated costs (3,4). These effects might be mediated by factors such as increased physiological strain, resource competition, modification of the gut flora and strain on organ systems (3).

Previous research on diarrheal comorbidity in children has largely focused on disease pair comparisons with prevalent conditions such as acute respiratory illness (5), malaria (6) and anemia (7). This approach, while valuable in understanding specific disease interactions, may not fully capture the complex landscape of multiple comorbidities often present in children with diarrhea. Furthermore, the landscape of childhood diarrheal illness has undergone significant transformations since 2008, driven by dynamic shifts in the epidemiological triad. Environmental factors, particularly those linked to ongoing climate change, have demonstrably altered the distribution of disease vectors (8) and potentially increased the incidence of diarrhea (9). At the host level, there has been reduction in HIV-infection and mother-to child transmission, stunting, and wasting (10,11). Additionally, targeted interventions such as the upscaling of zinc (12) and the introduction of rotavirus vaccines (13) have reshaped the management and prevention of diarrheal diseases. Concurrently, the evolution of pathogens at the agent level has introduced new challenges, most notably the rise of antimicrobial resistance (14) and serotype distribution (15). Moreover, several non-diarrhea-specific public health interventions and vaccination programs have been implemented. These include but are not limited to, RTS,S malaria vaccine, pneumococcal conjugate vaccine (PCV10), prevention of mother-to-child transmission initiatives, vitamin A supplementation, and distribution of insecticide-treated nets (ITNs). Understanding the patterns of diarrheal comorbidity and its associated factors against this backdrop of evolving epidemiological dynamics is imperative to inform the development of integrated strategies to effectively address this public health problem. Here, we leverage data from three consecutive enteric studies to explore temporal patterns and determine risk factors of diarrheal comorbidity among children aged < 5 years in rural western Kenya.

## Materials and Methods

### Study Design

This is a secondary, pooled individual-participant data analysis with a retrospective cohort design, using data from three prospective enteric multicenter studies, (i) the Global Enteric Multicenter Study (GEMS, 2008-2012), (ii) the Vaccine Impact on Diarrhea in Africa (VIDA, 2015-2018), and (iii) the Enteric for Global Health (EFGH) *Shigella* surveillance study (2022-2024).

### Study Setting and Population

The three studies were conducted in a predominantly rural area of western Kenya, previously described by Odhiambo et al. (16) and more recently by Omore et al. (17), with a population of 217,952 as of 2023. The population has a high level of primary education attainment (88.8%) and primarily engages in subsistence farming as the main economic activity. Vaccination coverage rates in the study area surpassed national estimates, with 88.1% of children aged 12-23 months having received core early childhood vaccines (One dose of Bacillus Calmette–Guérin (BCG) vaccine, three doses of the poliomyelitis vaccine, three doses of the diphtheria-pertussis-tetanus (DPT) vaccine, and one dose of the measles-mumps-rubella (MMR) vaccine). Additionally, 69.9% of the children were fully vaccinated according to the national immunization schedule (11). Siaya County still faces challenges of child malnutrition with 19.2%, 1.7% and 7.0% of children aged < 5 years reported to be stunted, wasted and under-weight, respectively. The reported HIV prevalence of 14.3% in the county is approximately 3 times the national average (18). In addition, the study area is situated within the lake endemic malaria zone, characterized by perennial intense malaria transmission and a prevalence rate of 27% (19).

### Data Sources

The three primary studies (GEMS, VIDA and EFGH) have been previously described (20–24). Briefly, GEMS was a 3-year prospective case-control study designed to assess the population-level burden, etiology, and clinical consequences of moderate-to-severe diarrhea (MSD) among children under five years of age in sub-Saharan Africa (SSA) and South Asia. Moderate-to-severe diarrhea (MSD) cases, defined as children aged 0-59 months presenting at a sentinel health center with diarrhea (defined as ≥ 3 looser-than-normal stools within 24 hours) that began within the past 7 days after ≥7 diarrhea-free days and had ≥1 of the following: sunken eyes, poor skin turgor, dysentery, required intravenous rehydration, or hospitalization. Diarrhea-free controls matched by age, gender and geographical location were enrolled within 14 days of case enrolment. Following GEMS, the VIDA study employed the same design and methods to evaluate diarrheal etiologies, assess rotavirus vaccine effectiveness, and measure the population-level impact of rotavirus vaccine introduction among children <5 years residing in censused populations in 3 SSA countries. More recently, EFGH was conducted, using cross-sectional and longitudinal study designs, to establish incidence and consequences of *Shigella* medically attended diarrhea (MAD) among children aged 6-35 months within 7 country sites in Africa, Asia, and Latin America. Eligible MAD cases were children aged 6-35 months presenting at a sentinel health center with diarrhea (defined as ≥ 3 looser-than-normal stools within 24 hours) that began within the past 7 days after ≥2 diarrhea-free days. In this analysis, we restricted EFGH data to children who met the GEMS/VIDA MSD criteria (diarrhea with dehydration, dysentery, or requiring hospitalization). The authors did not have access to any information that could identify individual participants during or after data collection.

In all the three studies, trained study personnel collected comprehensive data at enrollment, encompassing demographic information, illness history, anthropometric measurements, clinical features and the socio-economic characteristics of the enrolled children. Study clinicians made diagnoses based on clinical history and physical exam.

### Outcome variable

Comorbidity count was treated as a count outcome variable based on Integrated Management of Childhood Illnesses (IMCI) case definitions (25) and clinician diagnoses at enrolment. We assessed ten comorbidities that included: malaria, bacterial infection, pneumonia, severe acute malnutrition (SAM), meningitis, acute febrile illness (AFI), respiratory Illness (non-pneumonia), anemia, stunting and wasting. The detailed definitions of each comorbidity are shown in Table S1.

### Independent variables

A comprehensive list of demographic, clinical features, water and sanitation infrastructure, and the socio-economic characteristics were assessed as potential covariates. We used the *GEMS-modified Vesikari score system* (26), a 17-point scoring tool adapted from the original 20-point scale developed by Ruuska and Vesikari. The main difference is that vomiting is captured as a simple yes/no variable, rather than being assessed by frequency and duration (days). Additionally, a continuous variable representing time (year) and climate covariates (monthly rainfall and temperature) were also included. Monthly daytime land surface temperature (LSTD) and nighttime land surface temperature (LSTN) data were extracted from the Moderate Resolution Imaging Spectroradiometer (MODIS) (27). Monthly rainfall data were processed from the Climate Hazards Group InfraRED Precipitation with Station data (CHIRPS) (28) while historical daily near-surface air temperature data were extracted from the ERA5-Land and processed to a monthly scale (29). We also included climate variables lagged by up to three months.

### Statistical Analysis

We summarized categorical variables by generating frequency tables with corresponding percentages while continuous variables were summarized using the median and interquartile range (IQR) for each study and for the combined dataset.

We calculated the annual frequency and prevalence of children with 0, 1, 2, 3, and ≥4 comorbid conditions. Prevalence was computed by dividing the number of children in each comorbidity group by the total number of enrolled children per year. To evaluate temporal trends in the prevalence of individual comorbidities and any comorbidity, we conducted a Cochran-Armitage trend test. To assess how comorbidity burden varied by age and study period, we categorized children into age groups (0–11 months, 12–23 months, 24–59 months) and calculated the prevalence of each comorbidity category within these strata as well as overall. We explored temporal trends for 10 individual comorbidities and any comorbidity with comorbidity prevalence plotted over time with separate trajectories shown for each condition and study period. To evaluate patterns of co-occurrence among comorbidities, we generated an UpSet plot showing intersecting comorbidity profiles.

We fitted generalized estimating equations (GEE) with a Poisson distribution and a log link, to determine the risk factors of comorbidity count (30). Initially, bivariate models were fitted to assess the unadjusted association between each predictor variable. All variables were included in the multivariable model except the Vesikari score, as it is a composite of other predictors. Potential collinearity among the selected predictors was assessed using the Cramer’s V statistic. To account for within-subject correlation and potential heteroscedasticity, we used an exchangeable correlation structure and specified robust standard errors via the Huber-White sandwich estimator. We reported the incidence rate ratios (IRRs) and their 95% confidence intervals (CIs). To assess the adequacy of the Poisson model, we calculated the overdispersion statistic as the ratio of the Pearson chi-square statistic to the residual degrees of freedom.

A ratio substantially greater than 1 was considered evidence of overdispersion. In our analysis, no evidence of overdispersion was observed. All the statistical analyses were carried out using R software, version 4.4.1 (R Foundation for Statistical Computing, Vienna, Austria)

### Ethical Considerations

The protocols for GEMS, VIDA, and EFGH studies were reviewed and approved by the Scientific and Ethical Review Unit (SERU) of the Kenya Medical Research Institute (KEMRI) with the approvals granted under KEMRI SSC Protocol #1155, SERU#2996 and SERU#4362, respectively. Additionally, GEMS and VIDA obtained approval from the Institutional Review Board (IRB) of the University of Maryland, School of Medicine with approvals granted under UMB Protocol #H-28327 for GEMS and UMB Protocol #: HM-HP-00062472 for VIDA. Moreover, the IRB for the Centers for Disease Control and Prevention, Atlanta, GA, USA approved the VIDA protocol (reliance agreement 6729), and formally deferred its review of the GEMS protocol to the University of Maryland IRB (CDC Protocol # 5038). Prior to initiating any study procedures, written informed consent was obtained from each caregiver of all participating children across all three studies.

## Results

### Baseline characteristics

A total of 4,148 children with diarrhea were analyzed, drawn from the three studies: GEMS (n=1,778), VIDA (n=1,554), and EFGH (n=816). The cohort’s median age was 14 months (IQR: 8–24), and 44.6% (n=1,850) were female. The baseline characteristics of patients overall and stratified by study are shown in Table 1. Caregiver education below the primary level was reported for 33.8% (n=1,400) of participants overall, with a downward trend from 44.8% in GEMS to 9.7% in EFGH. Access to improved water and sanitation was reported for 65.6% (n=2,720) and 38.3% of households, respectively, with improved sanitation showing an increasing trend across studies. Clinically, the median diarrhea duration was five days (IQR: 3–8). Over half of the children had seven or more episodes of diarrhea (56.8%) and vomiting (52.3%, n=2,170), while 9.7% (n=401) required admission. Based on a 17-point modified vesikari score, disease severity was moderate overall (median: 10; IQR: 8–11), with VIDA study having the most severe episodes (median: 11; IQR: 9–12) compared to GEMS and EFGH (median: 9; IQR: 7–11).

**Table 1.**
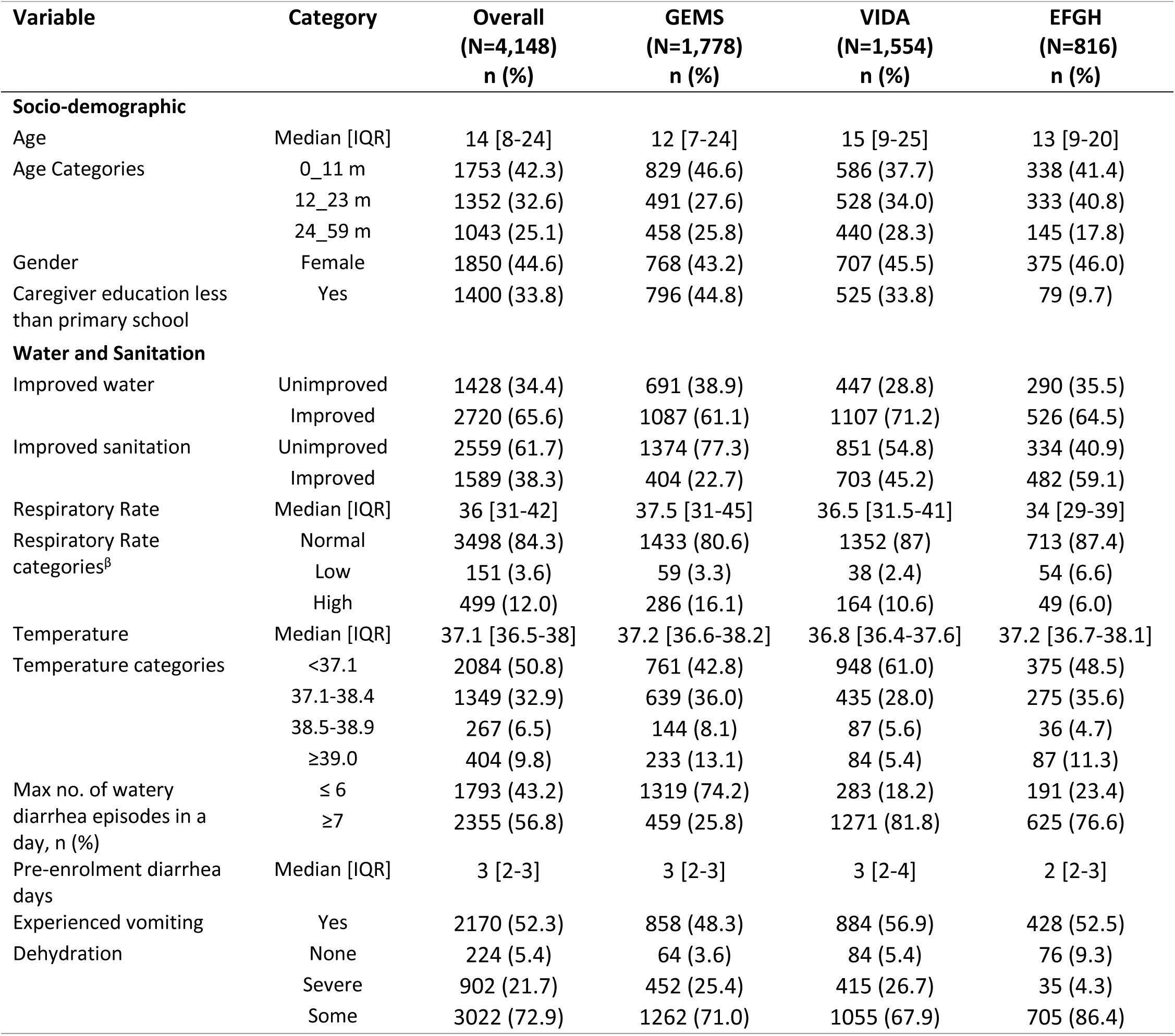
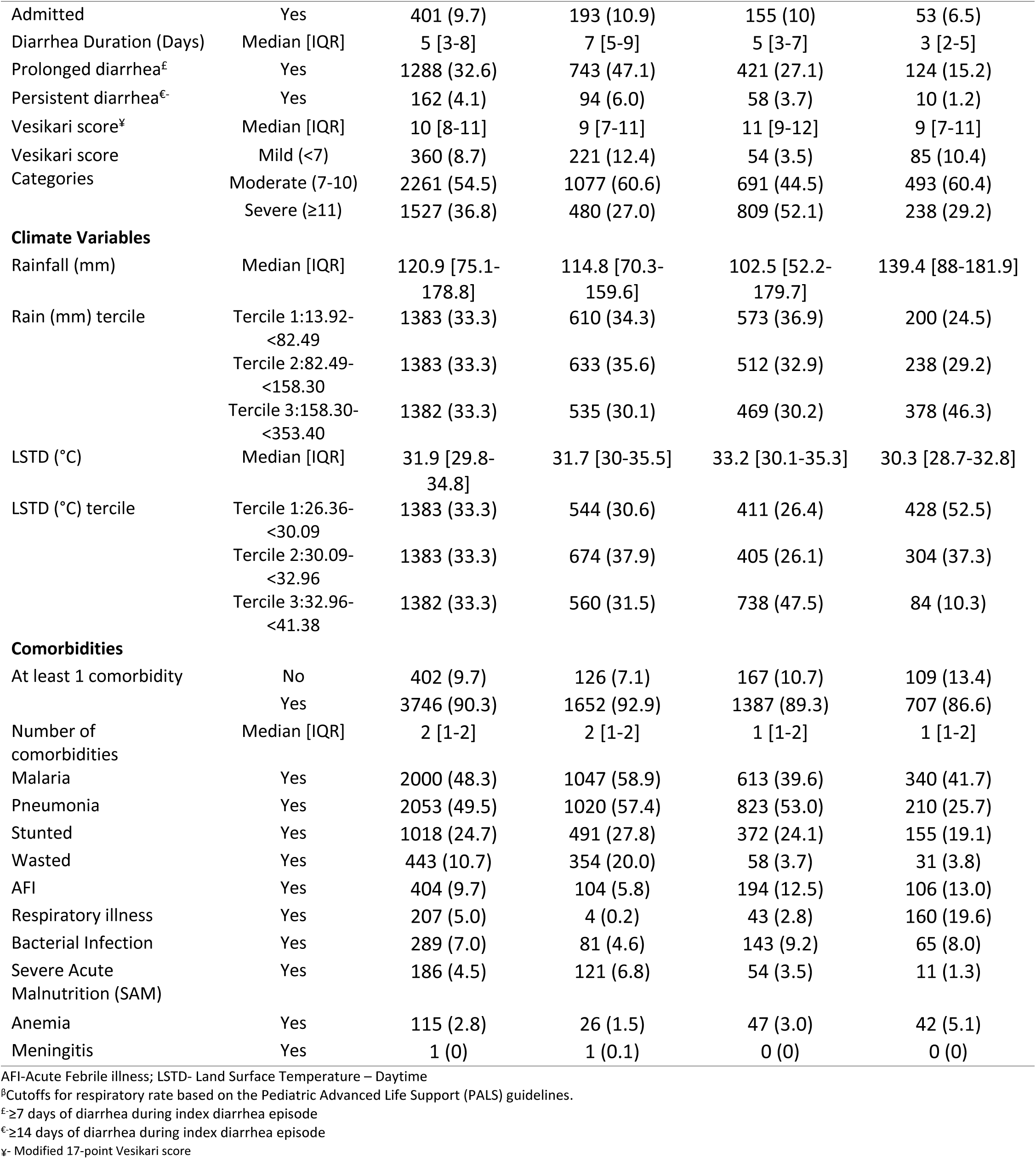
Characteristics of children aged < 59 months presenting with diarrhea in Western Kenya, 2008-2024.

### Patterns of Comorbidities

Majority of the children (90.3%, n = 3,746) had at least one comorbidity, with a declining trend across studies: GEMS (92.9%, n = 1,652), VIDA (89.3%, n = 1,387), and EFGH (86.6%, n = 707). The most common comorbidities were pneumonia (49.5%, n = 2,053), malaria (48.3%, n = 2,000), and stunting (24.7%, n = 1,018), with pneumonia and stunting declining over time across the three study periods (Table 1). Moreover, we observed distinct age-related patterns in comorbidity prevalence across the three study cohorts. In GEMS, which had the highest comorbidity burden, disease complexity peaked in younger children aged 12–23 months, who exhibited the highest prevalence of multiple comorbidities (≥2 [66.0%]). Conversely, the VIDA cohort had an intermediate burden and the EFGH cohort having the least comorbidity burden, and demonstrated a more linear trend. In VIDA the prevalence of having two or more comorbidities rose from 44% in the 0–11 month age group to 49% in the 24–59 month group. A similar, though less pronounced, increase was seen in the EFGH cohort, rising from 37% to 43% across the same age brackets (Fig 1).

**Fig 1.**
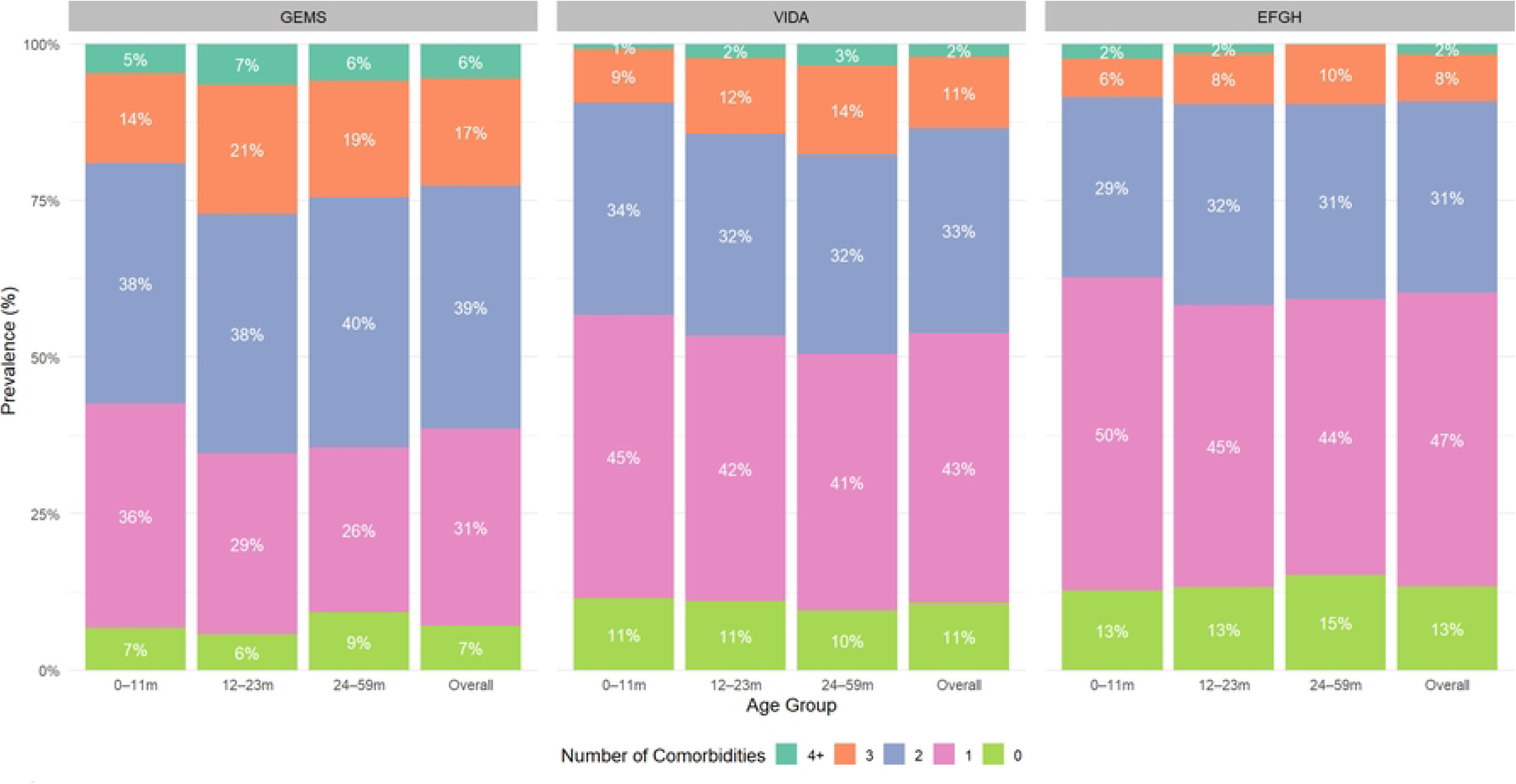
Age-stratified prevalence of number of comorbidities in childhood diarrhea in Western Kenya, 2008-2024

Over the 16-year period from 2008 to 2024, the burden of comorbidities has shown a shift towards fewer comorbidities per child, particularly a rise in children with exactly one comorbidity. The prevalence of patients having a single comorbidity increased, from 28.9% in 2008 to approximately half (49.7%) in 2024. Concurrently, there were declines in the prevalence of multiple comorbidities between 2008 and 2024, with reductions observed among children with two (38.8% to 30.1%), three (21.8% to 4.6%), and four or more comorbidities (7.7% to 2.0%). While the proportion with zero comorbidities showed some year-to-year fluctuation, but it generally increased compared to the earlier years (2.7% in 2008 vs. 13.7% in 2024) (Fig 2).

**Fig 2.**
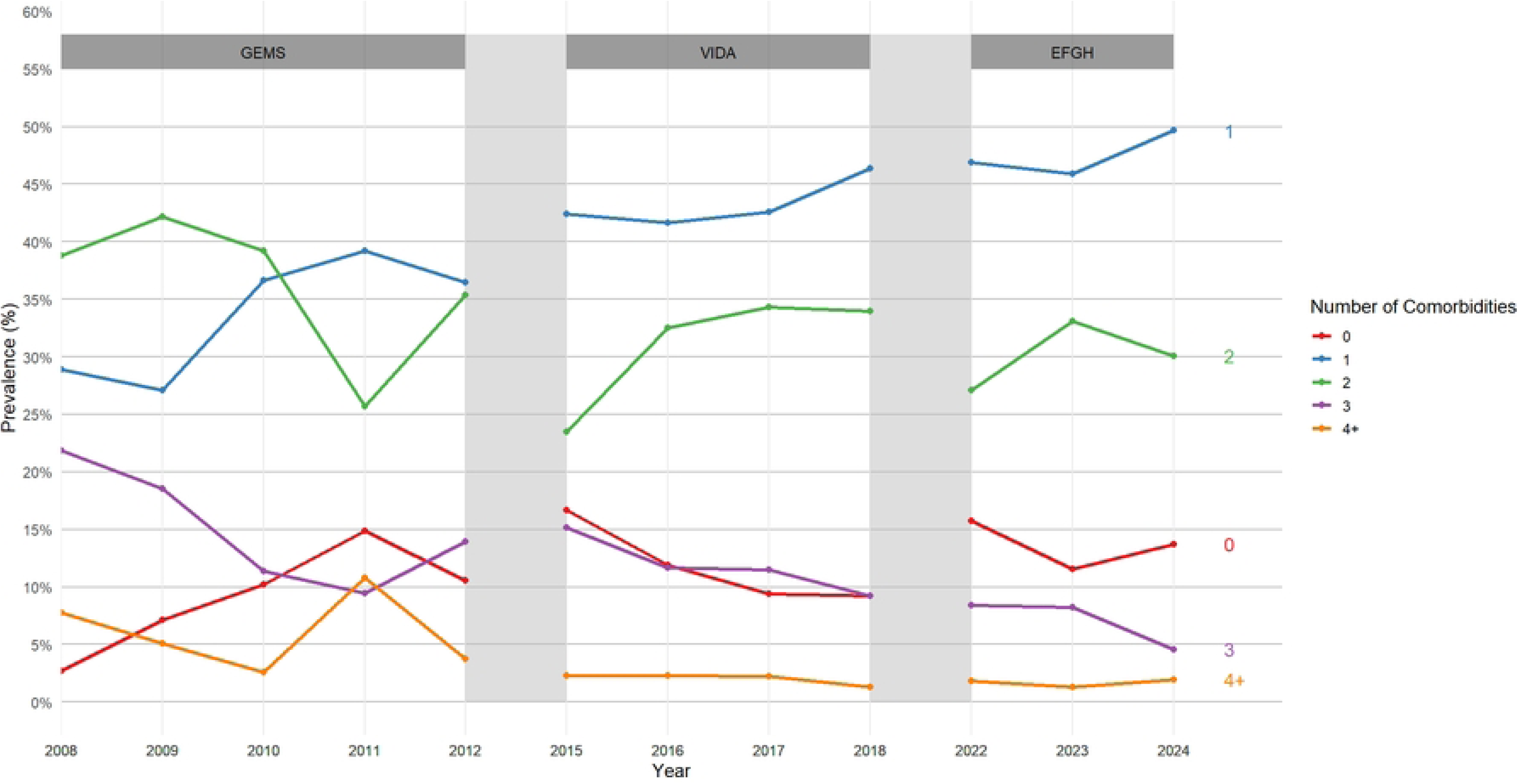
Prevalence of number of comorbidities in childhood diarrhea in Western Kenya, 2008-2024

During the study period, we observed a distinct epidemiological transition. Traditional major comorbidities like malaria, pneumonia, stunting, wasting, and SAM have shown significant reductions over the 16-year period (Fig 3; Table 2).

**Fig 3.**
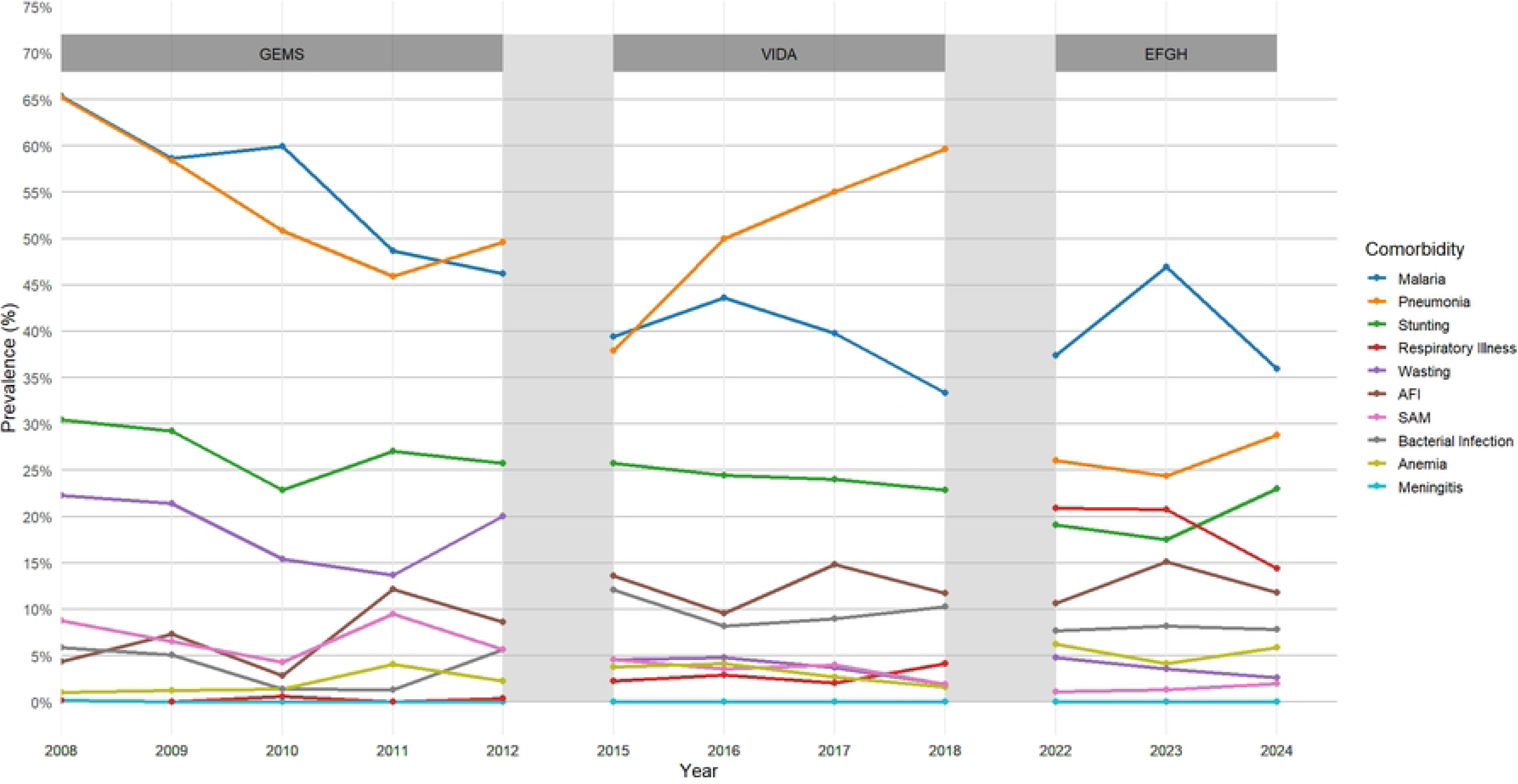
Prevalence of individual comorbidities in childhood diarrhea in Western Kenya, 2008-2024

**Table 2.** Temporal trends of diarrheal comorbidities among children aged <59 months in Western Kenya, 2008-2024.

| Comorbidity | 2008 | 2009 | 2010 | 2011 | 2012 | 2015 | 2016 | 2017 | 2018 | 2022 | 2023 | 2024 | p_value for Trend |
| --- | --- | --- | --- | --- | --- | --- | --- | --- | --- | --- | --- | --- | --- |
| Any Comorbidity | 579 (97.3%) | 456 (92.9%) | 316 (89.8%) | 63 (85.1%) | 238 (89.5%) | 110 (83.3%) | 423 (88.1%) | 568 (90.6%) | 286 (90.8%) | 230 (84.2%) | 345 (88.5%) | 132 (86.3%) | <0.001 |
| Malaria | 389 (65.4%) | 288 (58.7%) | 211 (59.9%) | 36 (48.6%) | 123 (46.2%) | 52 (39.4%) | 208 (43.6%) | 249 (39.8%) | 104 (33.3%) | 102 (37.4%) | 183 (46.9%) | 55 (35.9%) | <0.001 |
| Pneumonia | 388 (65.2%) | 287 (58.5%) | 179 (50.9%) | 34 (45.9%) | 132 (49.6%) | 50 (37.9%) | 240 (50.0%) | 345 (55.0%) | 188 (59.7%) | 71 (26%) | 95 (24.4%) | 44 (28.8%) | <0.001 |
| Stunting | 180 (30.5%) | 143 (29.2%) | 80 (22.9%) | 20 (27%) | 68 (25.8%) | 34 (25.8%) | 117 (24.5%) | 149 (24.0%) | 72 (22.9%) | 52 (19.1%) | 68 (17.5%) | 35 (23%) | <0.001 |
| Wasting | 132 (22.3%) | 105 (21.4%) | 54 (15.4%) | 10 (13.7%) | 53 (20.1%) | 6 (4.5%) | 23 (4.8%) | 23 (3.7%) | 6 (1.9%) | 13 (4.8%) | 14 (3.6%) | 4 (2.6%) | <0.001 |
| AFI | 26 (4.4%) | 36 (7.3%) | 10 (2.8%) | 9 (12.2%) | 23 (8.6%) | 18 (13.6%) | 46 (9.6%) | 93 (14.8%) | 37 (11.7%) | 29 (10.6%) | 59 (15.1%) | 18 (11.8%) | <0.001 |
| Respiratory Illness | 1 (0.2%) | 0 (0%) | 2 (0.6%) | 0 (0%) | 1 (0.4%) | 3 (2.3%) | 14 (2.9%) | 13 (2.1%) | 13 (4.1%) | 57 (20.9%) | 81 (20.8%) | 22 (14.4%) | <0.001 |
| Bacterial Infection | 35 (5.9%) | 25 (5.1%) | 5 (1.4%) | 1 (1.4%) | 15 (5.6%) | 16 (12.1%) | 39 (8.2%) | 56 (8.9%) | 32 (10.3%) | 21 (7.7%) | 32 (8.2%) | 12 (7.8%) | <0.001 |
| SAM | 52 (8.7%) | 32 (6.5%) | 15 (4.3%) | 7 (9.5%) | 15 (5.6%) | 6 (4.5%) | 17 (3.5%) | 25 (4.0%) | 6 (1.9%) | 3 (1.1%) | 5 (1.3%) | 3 (2.0%) | <0.001 |
| Anemia | 6 (1%) | 6 (1.2%) | 5 (1.4%) | 3 (4.1%) | 6 (2.3%) | 5 (3.8%) | 20 (4.2%) | 17 (2.7%) | 5 (1.6%) | 17 (6.2%) | 16 (4.1%) | 9 (5.9%) | <0.001 |
| Meningitis | 1 (0.2%) | 0 (0%) | 0 (0%) | 0 (0%) | 0 (0%) | 0 (0%) | 0 (0%) | 0 (0%) | 0 (0%) | 0 (0%) | 0 (0%) | 0 (0%) | -- |

AFI- Acute Febrile illness; SAM- Severe Acute Malnutrition

Specifically, malaria decreased by nearly half from 65.4% in 2008 to 35.9% in 2024, while pneumonia declined from 65.2% to 28.8% over the same period. Similarly, the prevalence of stunting dropped from 30.5% to 23.0%, and wasting showed an even steeper reduction from 22.3% to just 2.6%. Cases of SAM also declined steadily from 8.7% in 2008 to 2.0% in 2024. On the other hand, respiratory illness (non-pneumonia), which was nearly absent in 2008 (0.2%), rose to peaks in 2022-2023 (>20%) before slightly decreasing to 14.4% in 2022 emerging as a new concern or a less severe manifestation of the causes of pneumonia which has decreased. Acute febrile illness (AFI) increased from 4.4% to peaks around 2017–2023, before slightly decreasing to 11.8 % in 2024. Additionally, anemia also rose slightly from 1.0% in 2008 to 3.4% in 2024, with a peak of 6.2% in 2022. Bacterial infections showed fluctuating trends, ranging from 1.4% in 2010 to around 7–10% in more recent years. All observed trends were statistically significant (p < 0.01), indicating meaningful changes in the comorbidity profiles of children with diarrhea over time (Table 2).

### Co-occurrence

Single comorbidities were the most common patient groupings with pneumonia alone reported in 566 (15.1%), malaria alone in 561 (15.0%), AFI alone in 218 (5.8%), and stunting alone in 107 (2.9%) —these being the four most frequent single comorbidities (Fig 4). We observed 32 distinct comorbidity co-occurrences with most common combinations involving malaria, pneumonia and stunting. Specifically, malaria and pneumonia was the most common combination 644 (17.2%), followed by pneumonia and stunting 161 (4.3%), the triad of malaria, pneumonia and stunting 158 (4.2%), and malaria and stunting 138 (3.7%). Although stunting alone was not the most prevalent single comorbidity, it was a frequent component in the most common disease combinations, highlighting its central role in overall patient morbidity.

**Fig 4.**
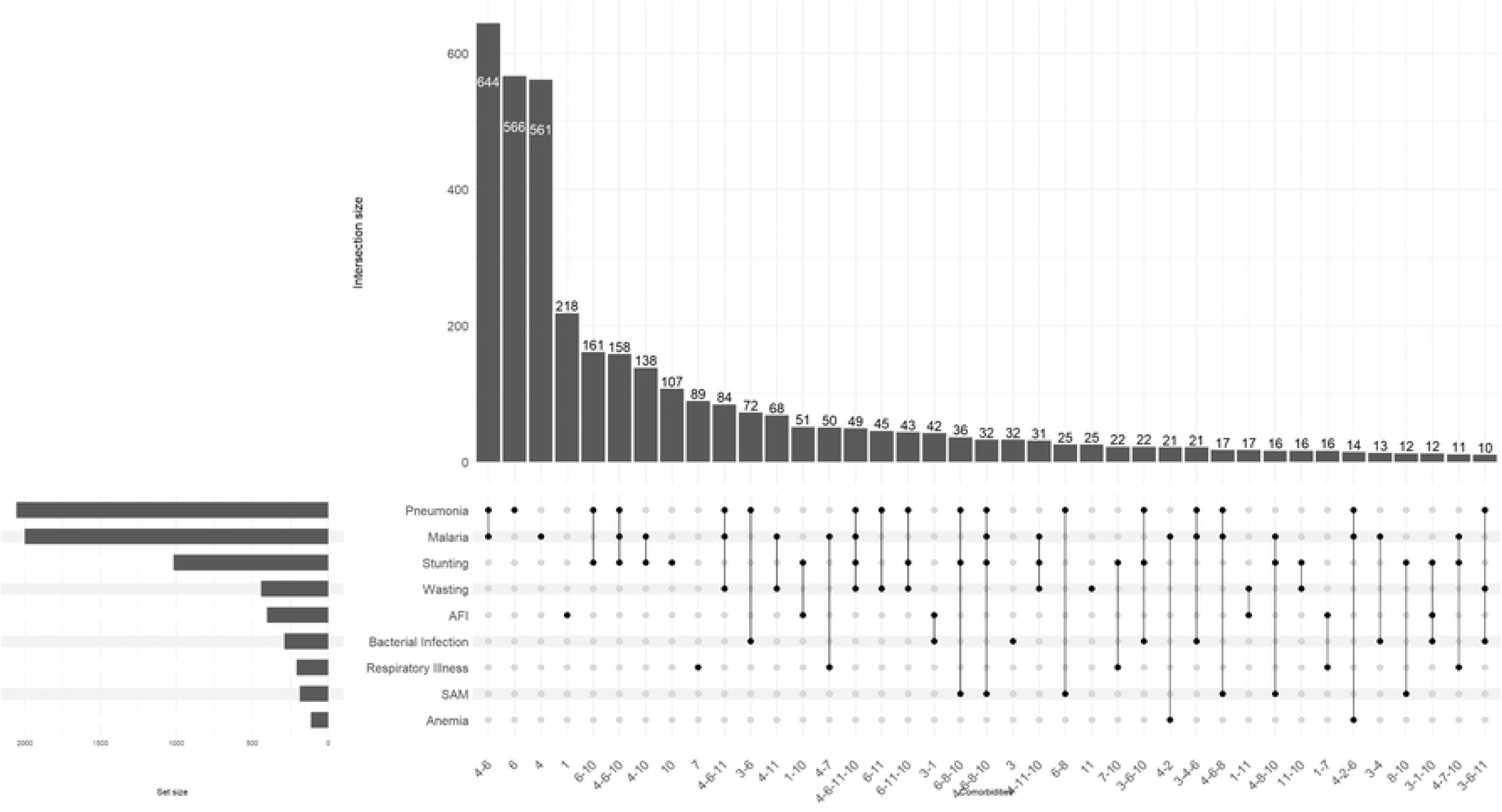
Upset plot of comorbidity co-occurrence in childhood diarrhea in Western Kenya, 2008-2024

### Risk Factors for Diarrheal Comorbidity

Based on our multivariable model, we observed that older children had a significantly higher number of comorbidities compared to infants 24–59 months: adjusted Incidence Rate Ratio (aIRR)= 1.10, 95% CI [1.05–1.14] (Table 3). Furthermore, children of caregivers having less than a primary school education had a 9 % higher number of comorbidities compared to those with caregivers having secondary or higher education (aIRR=1.09, 95%CI [1.05–1.13]). Similarly, dehydration was associated with 18% and 29% higher number of comorbidities for some dehydration (aIRR=1.18, 95% CI [1.07-1.30]) and severe dehydration (aIRR=1.29, 95%CI [1.16–1.43]), respectively. Other clinical signs associated with a significantly higher number of comorbidities included vomiting (aIRR: 1.25, 95% CI [1.21–1.29]) and a high respiratory rate (aIRR=1.22, 95%CI [1.16–1.27]). Finally, each additional day of diarrhea prior to enrolment was associated with a 2% higher number of comorbidities (aIRR: 1.02 [1.00–1.03]) (Table 3). While three-month lagged land surface temperatures were associated with 7% and 8% higher number of comorbidities in Terciles 2 and 3, respectively, at the bivariate level, this association was not retained in the multivariable model.

**Table 3.**
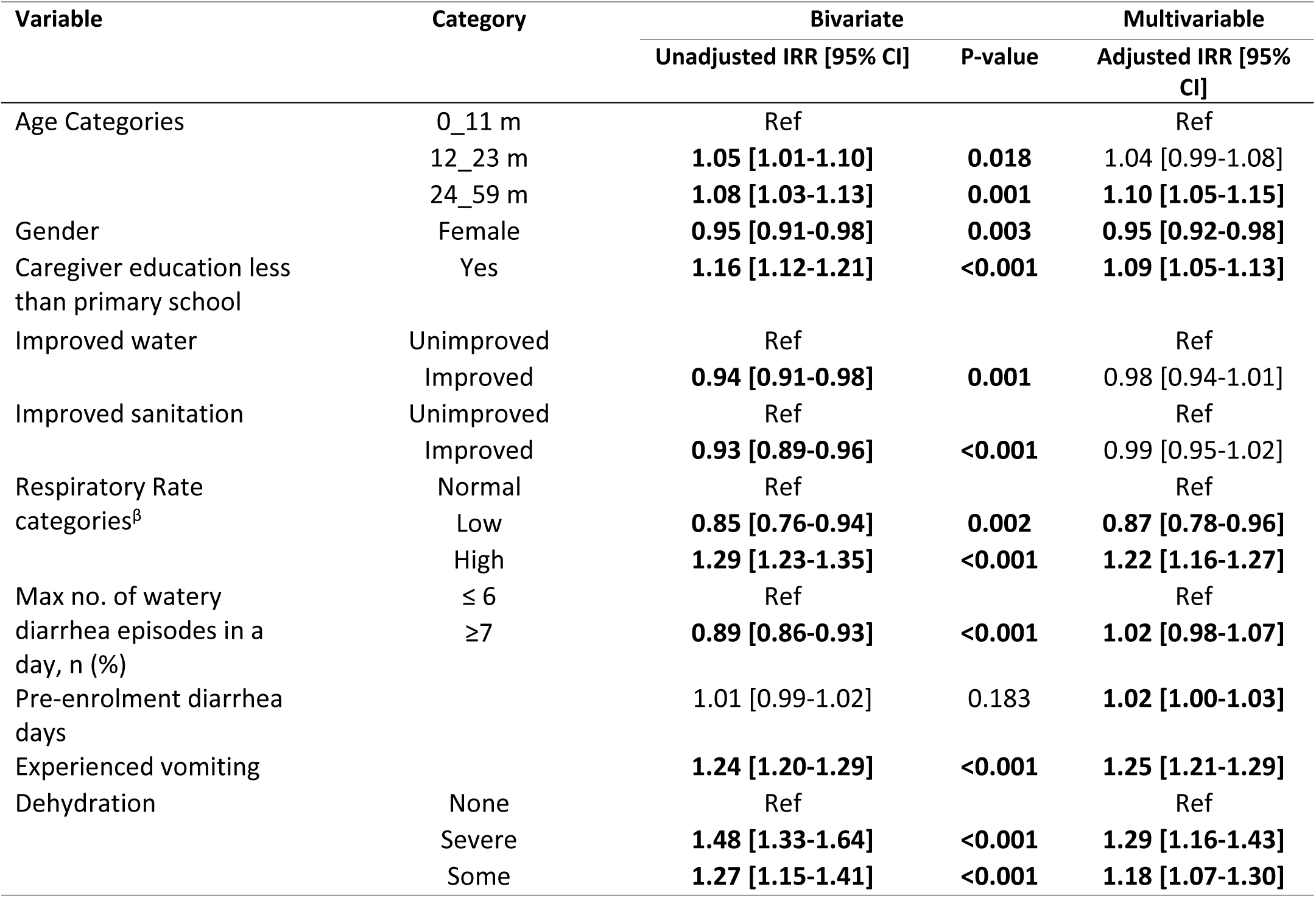
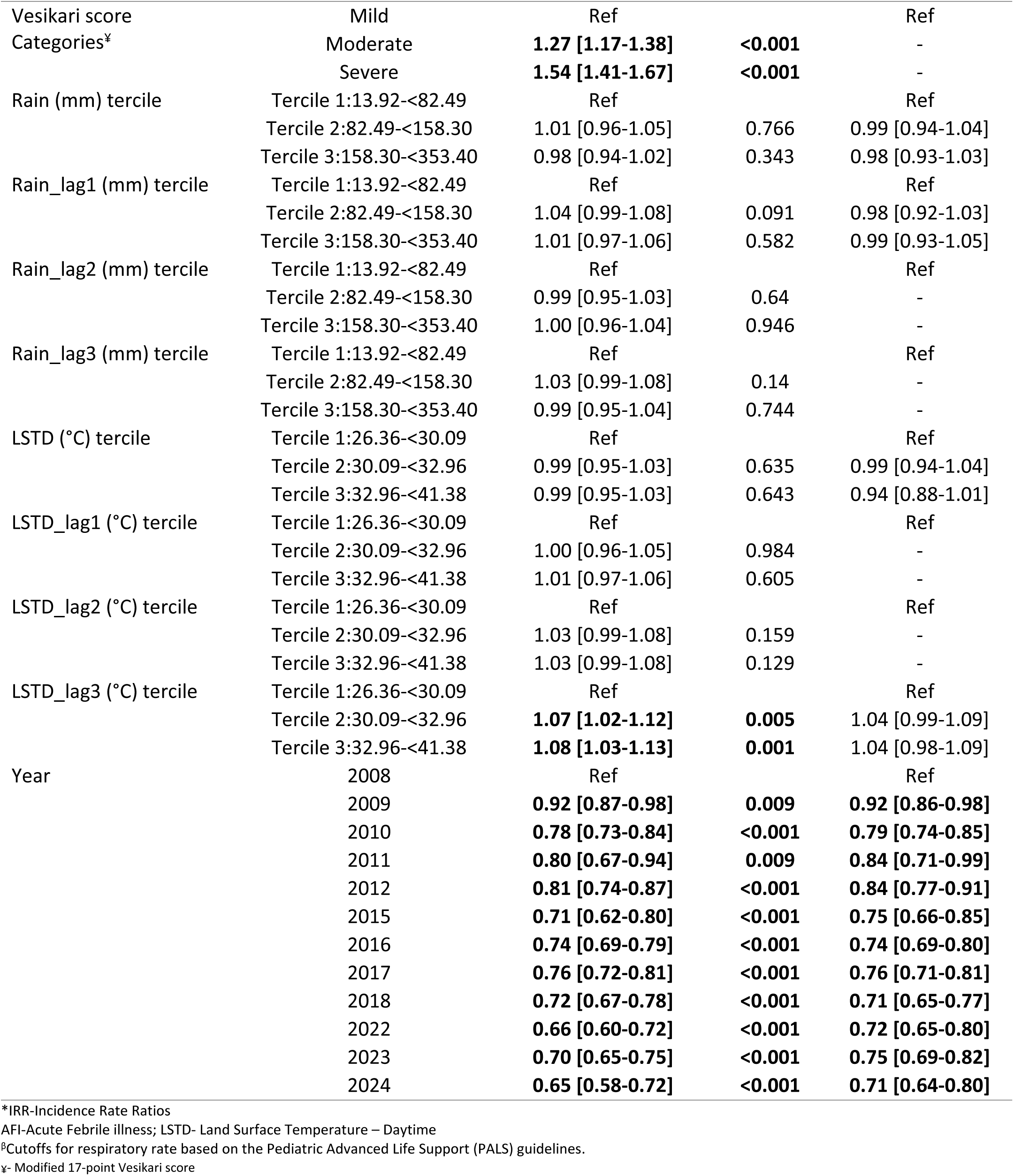
Factors associated with number of comorbidities in childhood diarrhea in Western Kenya, 2008-2024.

Conversely, female gender was associated with 5% fewer comorbidities compared to males (aIRR=0.95, 95%CI [0.92–0.98]). Importantly, there was a significant and consistent downward temporal trend across the study period. Compared to the reference year 2008, the rate of comorbidities decreased progressively over time, with the number in the most recent years (2022–2024) being approximately 25-29% lower (2022: aIRR= 0.72, 95% CI [0.65–0.80]; 2023: aIRR=0.75, 95% CI [0.69–0.82], 2024: aIRR=0.71, 95% CI [0.64–0.80]) (Table 3).

## Discussion

Over the 16-year period (2008–2024), the burden and profile of comorbidities observed among children with diarrhea in Kenya has shifted significantly. While comorbidities remain common (90.3% overall), there is a clear decline in both prevalence and severity over time and across successive studies. Pneumonia, malaria, and stunting were the most prevalent single comorbidities as well as the most common co-occurrence combinations. Additionally, there has been a shift toward fewer comorbidities per child, with an increase in children presenting with only one comorbidity and a decline in those with multiple conditions (≥ 2 comorbidities). Moreover, traditional comorbidities (malaria, pneumonia, wasting, and SAM) have declined significantly, while respiratory illness (non-pneumonia), AFI, and anemia have emerged or increased. Stunting frequently co-occurred with other conditions, underlining its role in compounded morbidity. Finally, multivariable analysis identified older age, lower caregiver education, dehydration, vomiting, and high respiratory rate as significant predictors of higher comorbidity counts, while female sex was associated with fewer comorbidities. Importantly, comorbidity burden decreased significantly over time, with 25–29% fewer comorbidities in recent years (2022-2024), reflecting a broader epidemiological transition.

The observed decline in the prevalence and severity of diarrheal comorbidities over time may reflect broader improvements in child health, nutrition, and healthcare systems. The likely most important driver of this observation is the success of targeted public health interventions and vaccination programs that directly target the most common comorbidities observed. Furthermore, reduced mother-to-child transmission of HIV in this setting may also explain in part the reduction of the burden of diarrheal comorbidity observed. Moreover, the scale-up of prevention of mother-to-child transmission (PMTCT) programs in Kenya (over 90% coverage) has likely contributed to the decline in diarrheal comorbidity by reducing pediatric HIV infections and related immunosuppression, which increases vulnerability to severe diarrhea and other opportunistic infections (10). This decline may have been further accentuated by the introduction and scaling up of Vitamin A supplementation through the Child Health Weeks campaign, launched in 2007. Increased coverage over time likely enhanced immune function, contributing to reduced severity and duration of diarrheal episodes and associated complications (31). However, a paradox of this public health success combined with evolving diagnostic practices and underlying environmental factors is the emergence of a secondary layer of health issues that were previously overshadowed or misdiagnosed. Respiratory illness (non-pneumonia), AFI, and anemia have gained relative importance in the overall comorbidity burden of childhood diarrhea even as traditional comorbidities (malaria, pneumonia, wasting, and SAM) have declined.

Specifically, the introduction of the rotavirus vaccine in 2014 has reduced the incidence and severity of viral diarrhea (32,33). The residual viral diarrheal episodes are likely less severe leading lower risk of severe dehydration, thereby reducing associated comorbidities. Furthermore, the updating of national guidelines for pediatric diarrhea to include zinc in 2007 alongside Oral rehydration solution (ORS) has worked to shorten duration and reduce severity of diarrhea, potentially lowering the risk of secondary complications (34). In parallel, the introduction of the Pneumococcal Conjugate Vaccine (PCV10) into the national immunization program in 2011 has led to substantial reduction in PCV10-type pneumonia cases possibly explaining the reduced prevalence of pneumonia as a comorbidity (35). This decline has been matched with an increase in respiratory illness (non-pneumonia), which likely reflects the persistent, and perhaps increasing, circulation of viral agents like Respiratory Syncytial Virus (RSV), influenza, parainfluenza, and more recently, SARS-CoV-2 (36,37). Additionally, urbanization, increased population density, and environmental changes including climate variability may be facilitating the spread of respiratory viruses (38). The distinct peaks in respiratory illness observed in 2022-2023 may also reflect post-COVID-19 lockdown surges in common viral pathogens as population immunity shifted. Similarly, gradual reductions in childhood stunting have also been reported in the country, with a reduction rate of 1.6% per annum reported between 1993-2014 largely driven by improved national socio-economic well-being (39). The most recent 2022 Kenya Demographic Health Survey reported a prevalence of 18.0% (11).

In synchrony, the significant drop in malaria as a comorbidity directly correlates with the nationwide scale-up of insecticide-treated nets (ITNs) and effective artemisinin-based combination therapy (ACT), which began in 2006 (40). This reduction in malaria prevalence has also been heightened by the pilot introduction of the RTS,S malaria vaccine in high-burden counties including Siaya County in 2019 (41). Additionally, the widespread use of Rapid Diagnostic Tests (RDTs) and microscopy has reduced the over-diagnosis of malaria (42), directly explaining the rise in AFI, which is often a diagnosis of exclusion, capturing a wide range of viral (including dengue, chikunguya) and bacterial infections (salmonellosis, rickettsia). Of note, primary healthcare settings lack the capacity to diagnose the etiology of AFI beyond malaria testing, leading to this broad category gaining prominence. Additionally, the emergence of anemia is consistent with findings from Hailu et al., which showed an increase in the prevalence of anemia in the East Africa region in the period 2016-2021 following gradual declines in 2006-2015 (43). This could possibly be explained by the fact that despite the decline in malaria, malaria remains endemic in the study area possibly causing hemolytic anemia in young children (19). Additionally, anemia is closely linked to poor nutrition, with stunted and wasted children more affected than their peers possibly caused by inadequate maternal nutrition, and low intake of iron-rich foods (44).The impact of the interventions and vaccinations programs discussed above may explain in part the decline in prevalence of the leading comorbidities as well as the overall burden of comorbidity count in childhood diarrhea.

The higher comorbidity count in older children compared to younger children likely reflects increased environmental exposure due to greater mobility and social interaction (45), combined with a maturing immune system that is still developing after the loss of maternal antibodies (46), making them more vulnerable to overlapping infections. Moreover, lower caregiver education is associated with higher comorbidity in children likely due to limited health knowledge, delayed care-seeking, poor hygiene and nutrition practices, and overall socioeconomic disadvantage (47). These factors increase children’s exposure to infections and reduce their chances of timely treatment, leading to a higher risk of multiple, overlapping illnesses. Dehydration, vomiting, and high respiratory rate are predictors of higher comorbidity count because they signal severe physiological disruption, creating a cascade of vulnerability. Dehydration strains body organs and disrupts metabolism, leading to acidosis and tissue hypoxia, which weakens defenses and enables secondary infections (48). Vomiting prevents rehydration and nutrition, causing electrolyte imbalances, catabolism, and immunosuppression. Similarly, a high respiratory rate serves as a dual indicator, pointing either directly to a pulmonary comorbidity or to the body’s attempt to compensate for metabolic acidosis driven by the combined severity of multiple illnesses (49). Collectively, these signs signify a cycle of organ dysfunction and immune compromise that elevates susceptibility to secondary infections, thereby increasing the total comorbidity burden.

Conversely, females were more likely to have fewer comorbidities, a finding that is corroborated by Muenchhoff et al., who observed a strong sexual dimorphism in childhood infection outcomes, with males often more susceptible to many pathogens (50). This may be partly due to females exhibiting stronger Th1 immune responses, even before puberty and full expression of sexual traits.

Our findings highlight the critical need for sustained investment in interventions targeting traditional comorbidities including vaccination, malaria control, PMTCT of HIV, and nutrition initiatives to maintain the successes witnessed. However, there is need for public health priorities to pivot to address emerging comorbidities (respiratory illness [non-pneumonia], AFI, and anemia) by tackling the multifaceted drivers of anemia, respiratory risk factors, and improving diagnostics and management for AFI. Moreover, the frequent co-occurrence of stunting with infectious diseases demonstrates that siloed programs are insufficient. Our findings advocate for integrated care packages that combine nutrition support, vaccination, WASH promotion, and infectious disease management at every point of contact with the health system.

Our study is not without limitations. First, reliance on clinician diagnoses for comorbidities introduces potential subjectivity and misclassification bias. Second, the temporal sequence of co-occurring conditions is unclear: whether they developed concurrently or sequentially. Third, treating all comorbidities equally overlooks clinical and temporal differences between chronic and acute conditions. Fourth, restricting the analysis to one Kenyan site may limit generalizability of findings to other settings with different epidemiological or socio-economic profiles. The 16-year timeframe also introduces potential for unmeasured temporal confounders such as evolving healthcare access or public health interventions. Lastly, despite covariate adjustment, unmeasured factors like care-seeking behavior or underlying immunodeficiency may still confound associations.

## Conclusion

Over the past 16 years, Kenya has achieved a remarkable epidemiological transition in childhood diarrheal morbidity, marked by a 25–30% decline in comorbidity burden, despite the high prevalence, and a fundamental shift in disease profiles. The reduction in severe traditional comorbidities (malaria, pneumonia, wasting, and stunting) stands as a testament to the success of scaled interventions. However, this progress has unmasked a new challenge: the rising relative burden of anemia, respiratory illness (non-pneumonia), and AFI, now dominant features of the comorbidity landscape. Our findings serve as a call to action for Kenya’s strategy to evolve from single-disease control to integrated comorbidity management.

## Declarations

## Acknowledgements

We extend our deep gratitude to the families who participated in the three studies, and to the clinical and field teams whose dedication and hard work made this research possible. We are also grateful to the physicians, administrators, and Ministry of Health officials at each recruitment center in Siaya County for their provision of facilities, operational support, and gatekeeper permissions that enabled the successful implementation of the studies.

## Data Availability

The GEMS and VIDA datasets are publicly available on ClinEpiDB (https://clinepidb.org/ce/app) while the EFGH data will be made publicly available through the Vivli data-sharing platform in the near future.

